# An 8-DNA methylation signature predicts recurrence risk of cervical cancer

**DOI:** 10.1101/2021.04.29.21256287

**Authors:** Jing-Hang Ma, Yu Huang, Lu-Yao Liu, Zhen Feng

## Abstract

**Purpose:** Cervical cancer is the leading cause of cancer-associated mortalities in female worldwide. DNA methylation have been demonstrated to have a regulatory role in the progression and could be novel biomarkers to predict the recurrence of the disease.

**Methods:** DNA methylation and RNA expression data of cervical cancer were downloaded from TCGA. DMGs and DEGs were screened and extracted by correlation analysis. The SVM-based recurrence prediction model was established using the selected DMGs. Cox regression analysis and ROC analysis were used as self-evaluations. The GEO database was applied for external validation. Functional enrichment was determined by GO and KEGG analysis.

**Results:** The eight-DNA methylation signature could identify patients with a high risk of recurrence (AUC=0.833). The SVM score was an independent risk factor for recurrence (HR = 0.418; 95% CI: 0.26-0.67, p-value <0.05). The independent GEO database further supported the result.

**Conclusion:** The predictive function of the 8-DNA methylation signature for recurrence of cervical cancer was revealed in this study, which may help identify high-risk patients of recurrence and benefit the clinical treatment.

## Introduction

Cervical cancer is the leading cause of cancer-related deaths in women, with more than 500,000 patients diagnosed and 260,000 deaths each year worldwide [1]. Surgery is the main treatment for early-stage cervical cancer patients. For those with a high risk of recurrence, radiotherapy alone or in combination with chemotherapy is also needed [2]. It has been reported that the recurrence rate of cervical cancer is 20%–30%, which increases with disease stage. Recurrent patients always have poor prognosis, and therapeutic methods are limited with low response rate[3–5].

At present, the staging system (FIGO stage or TNM stage) together with the “Sedlis Criteria” and other risk factors are generally used to identify cervical cancer patients with a high risk of recurrence according to clinical and pathological characteristics to guide adjuvant treatment [6, 7]. These assessments of recurrence risk undoubtedly provide predictive value; however, they are not as good at determining cancer recurrence in comparison with genetic, epigenetic and molecular biomarkers. Therefore, it is necessary to identify biomarkers for predicting recurrence risk and determining treatment strategies to improve patients’ prognoses [8, 9].

DNA methylation is responsible for regulating gene transcription without changing the DNA sequence. Abnormal hypermethylation of tumor suppressor genes and hypomethylation of oncogenes are vital during the early tumorigenesis process [10, 11] and related epigenetic drugs have been proved to be effective in the treatment of cervical cancer[12]. Compared to the complex variability in RNA expression between individual patients [13], DNA methylation is a relatively stable and pervasive modification across tumor types that is associated with the pathogenesis and progression of cancer and related to the prognosis and outcome of patients [13]. Researchers have explored the relation between DNA methylation and cervical cancer, including the biological function of genes [14–16], methylation-based tumor classification [17, 18] and radiotherapy-related predictors [19]. However, few studies have investigated a recurrence prediction model of DNA methylation in cervical cancer, which is of clinical significance to help clinicians find high-risk patients and implement active treatment.

In this study, we used DNA methylation and related RNA expression datasets of cervical cancer patients from The Cancer Genome Atlas (TCGA) to explore the epigenetic signature for recurrence risk prediction.

## Methods

### Sample selection and data processing

DNA methylation datasets, RNA expression datasets and clinical information of cervical cancer patients used for modeling were downloaded from the TCGA (https://portal.gdc.cancer.gov/) [20] with the use of the FireBrowse Data Portal (http://firebrowse.org/) [21]. Illumina Infinium HumanMethylation450 BeadChip sequencing technology was used to obtain 312 DNA methylation profiles, and Illumina HiSeq2000 was used for 307 mRNA-seq profiles.

All samples in the TCGA were collected from primary tumors of cervical tissue, with tumor purity ranging from 22% to 96% [20]. We excluded samples with missing survival information and included samples according to the recurrence status and disease-free survival time. After screening and filtration, 85 patients who had been followed-up for more than two years without recurrence were included in the no recurrence group, and 50 patients with evidence of recurrence were included in the recurrence group, as recurrence usually occurs in the first two years after completing treatment [22, 23].

### Differential methylation analysis and differential expression analysis

DNA methylation datasets and RNA expression datasets from the TCGA were analyzed in R (version 3.6.0). DNA methylation levels (β values) with a p-value less than 0.01 and with an absolute log fold change larger than 0.025 were considered to be differentially methylated genes (DMGs). RNA expression levels with a p-value less than 0.01 and an absolute log fold change larger than 0.4 were considered to be differentially expressed genes (DEGs). The log fold change thresholds were set to identify genes proximately ranked in the top 1%. DMGs overlapping with DEGs were extracted for the correlation analysis. We used the *lm* function in R to fit the linear model. Those with a significant negative correlation between DMGs and DEGs were further chosen to establish the SVM-based prediction model.

### SVM model

SVM is a machine learning algorithm that offers a direct approach to binary classification [26]. The SVM considers data as points in a high-dimensional space and finds an optimal hyperplane to separate data into classes.

The eight selected DMGs from 135 patients in the TCGA database were denoted as 135 pairs (x_1_, y_1_), (x_2_, y_2_), …, (x_135_, y_135_). x_i_∈R^8^: eight-dimensional data represent methylation levels of eight DMGs in the i^th^ sample; y_i_∈{-1,1}: labels of classification. (x_i_, y_i_) were mapped into data points in the eight-dimensional space and were used to train the SVM-based prediction model without cross validation. The model found the optimal linear hyperplane P0{x: φ(x)= 0} in the feature space, where φ was the linear transformation. Points satisfying {x: φ(x)>0} or {x: φ(x)<0} belonged to the class y=1 or y= −1, respectively (the graphical illustration was shown in Figure.3b).

We extracted the φ function along with its coefficients from the linear SVM model and generated the SVM score to predict the classification. The SVM score was calculated with the β values of the eight DNA methylation genes and their coefficients in the trained SVM model. A constant bias was also introduced. A sample was classified into the low-risk or high-risk class if the SVM score was higher or lower than the threshold 0. In this work, we used the e1071 package in R [27], which contained the *svm* function.

### Recurrent signature validation

The confusion matrix was constructed to further confirm the prediction of the SVM classifier model. The ROC curve and the area under the curve (AUC) were used to assess the fit of the SVM-based prediction model [28]. A T-test and Fisher’s exact test were used to identify clinical features affecting recurrence. The terms with p-value<0.05 were retained for further multivariate Cox regression analysis.

### Data visualization using t-SNE

The t-distributed Stochastic Neighbor Embedding (t-SNE) algorithm [29], a nonlinear dimensionality reduction algorithm, was used to keep the topological structure in the high dimension and maps each datum onto several different, but related, low-dimensional manifolds, since an SVM plot can not visualize the eight-dimensional data of the eight DNA methylation genes.

We used the Rtsne package in R [30]. The parameter “perplexity”, balanced attention between local and global aspects of the data and was set to 30, which was an estimate of the approximately 30 closest neighbors each data point had. The number of iterations was 10,000.

### External validation of the GEO database

We searched cervical cancer datasets containing both DNA methylation and survival information across the GEO database, and the only dataset GSE30759 (the Illumina Infinium 27k Human Methylation BeadChip for DNA methylation profiles) with 63 samples of primary cervical cancer tissue satisfied such conditions. We performed a similar preprocessing on them as that used on the TCGA datasets. A patient whose recurrence-free survival time was equal to or longer than two years was regarded as a nonrecurrent patient. A patient whose survival time was shorter than the recurrence-free survival time was considered to suffer from the recurrence of the cancer.

The recurrence-related methylation genes in the GEO dataset overlapped with the 8 reference genes obtained from the training model using the TCGA dataset. Three genes, *LOC100132215, TIGIT*, and *ZFYVE21*, in the GEO dataset were found to not have probes. We calculated the average DNA methylation levels of these three genes in the TCGA dataset and subtracted them from the SVM score threshold according to (eq. 2), eventually generating the reduced SVM score threshold.

We analyzed the distribution of the methylation levels for the other five genes in both datasets. We found four genes whose distributions in both datasets were similar. We then normalized the levels in the GEO dataset to those in the TCGA datasets to avoid bias.Normalization of the methylation value of genes from the *i*^th^ sample in the GEO dataset is as follows:

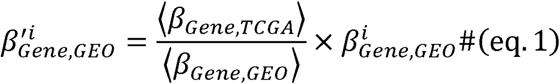

Of the five genes, *MPHOSPH10* was excluded due to the large differences in distribution between two datasets. Similarly, we subtracted the average DNA methylation level of the gene *MPHOSPH10* in the TCGA dataset to further update the reduced SVM score threshold.

### Functional enrichment analysis

Gene Ontology (GO) enrichment analysis [24] was performed to identify the function of DMGs, including biological process (BP), cellular component (CC) and molecular function (MF). Kyoto Encyclopedia of Genes and Genomes (KEGG) enrichment analysis was performed to identify KEGG pathways. The Database for Annotation, Visualization and Integrated Discovery (DAVID) was used to perform GO enrichment analysis and KEGG enrichment analysis (https://david.ncifcrf.gov/) [25]. The enrichment functions and pathways were considered significant if the p-value was < 0.05.

## Results

The study procedures are shown in **Figure 1**.

**Figure 1.**
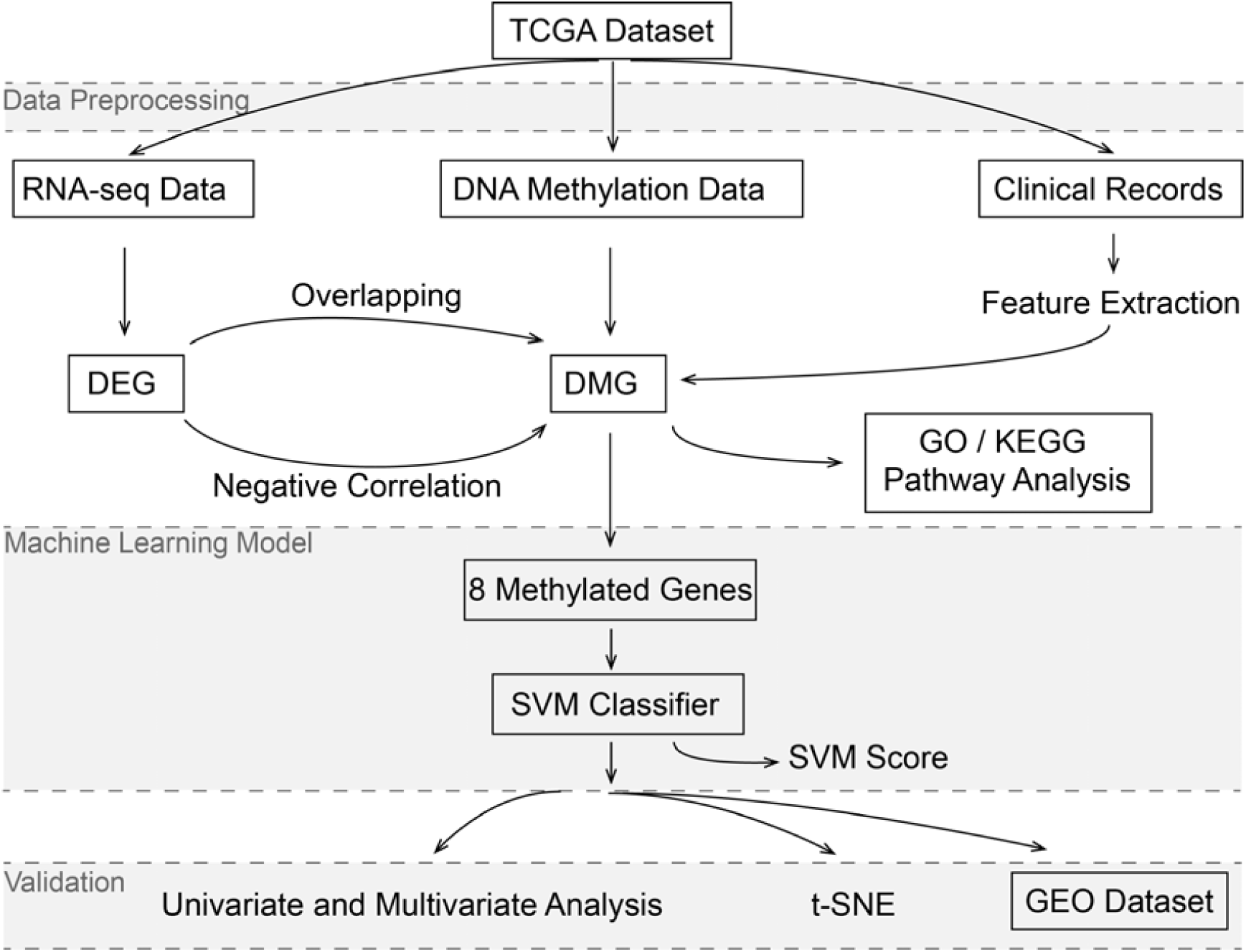
Flowchart of the whole analysis process.

### Identification of DMGs and DEGs

To identify the DMGs and DEGs between the recurrence and no recurrence groups, we screened 24,278 DNA methylation genes and 20,104 RNA expression data from the TCGA database (**Figure 2a**). Volcano plots were employed to show the distribution of DNA methylation or gene expression fold changes in each group (**Figure 2b, c**). A total of 305 DMGs (p-value <0.01; absolute log fold changes>0.025) and 262 DEGs (p-value <0.01; absolute log fold change value >0.4) were extracted. Eleven hypermethylated genes and 5 hypomethylated genes overlapped with the DEGs, and 8 of them (*ZFYVE21, C17orf62, PDIA6, CUL7, TIGIT, MPHOSPH10, LOC100132215* and *CD3E*) had negative correlations between DNA methylation and mRNA expression levels, with p-values <0.05, as shown in **Figure 2d, e**. These 8 DNA methylation genes were used to further establish the recurrence prediction model.

**Figure 2:**
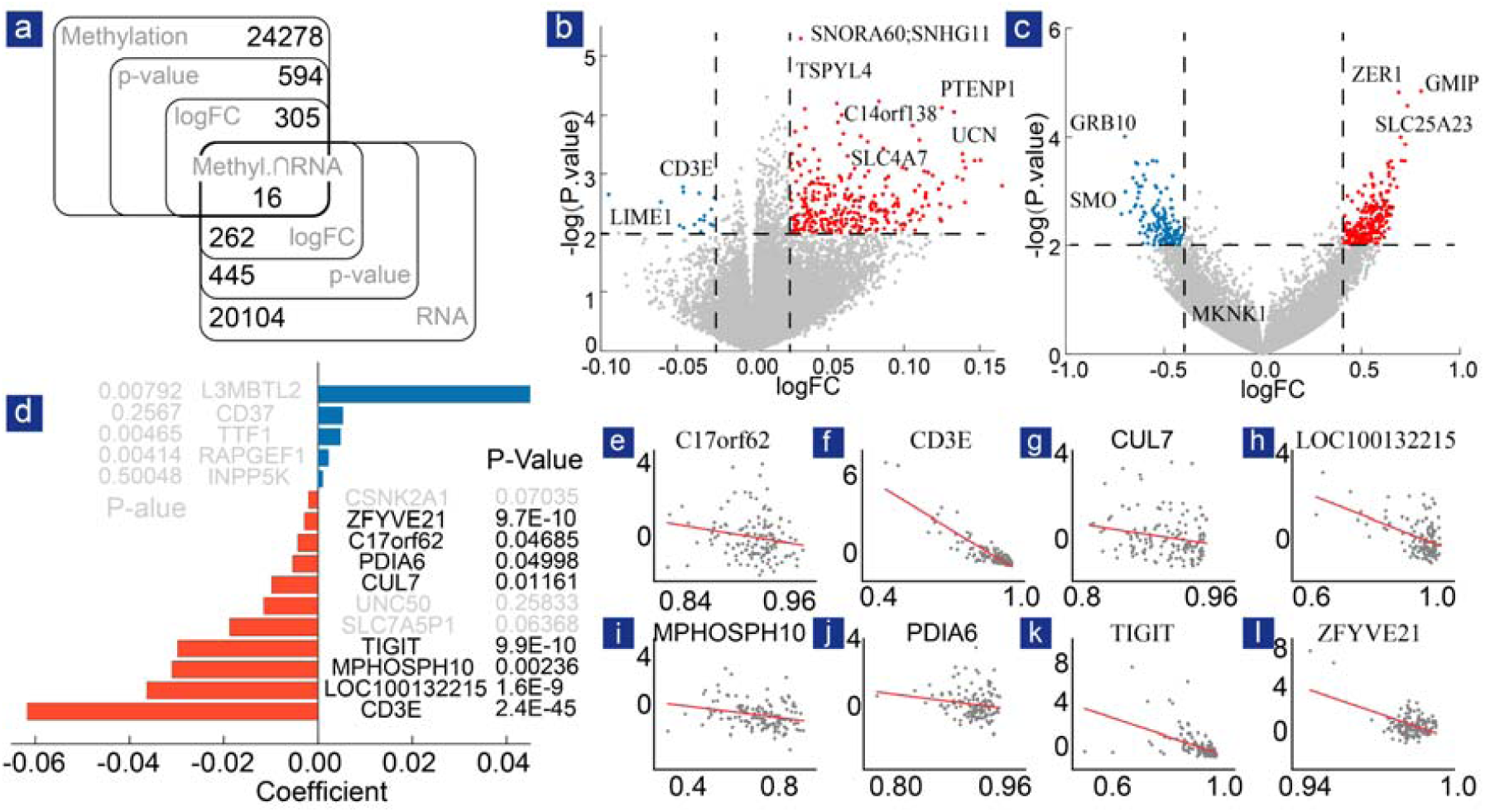
Selection of DMGs in recurrent and nonrecurrent cervical cancer samples. (a) Venn diagram of overlapping DEGs and DMGs. (b) Volcano plot of DMGs. (c) Volcano plot of DEGs. (d) Coefficient distribution of the gene signatures. (e) The correlation between gene expression and DNA methylation level in cervical cancer.

### The SVM-based prediction model of DNA methylation

The β values of the eight selected DNA methylation sites from the 135 patients in the TCGA dataset were visualized by a heat map using the pheatmap package in R (**Figure 3a**). We trained the SVM model according to the recurrence outcomes (**Figure 3b**). The confusion matrix is shown in **Figure 3c**, and the ROC curve (AUC: 0.833) is shown in **Figure 3d**. The SVM score was generated to predict the recurrence rate with the β values of the eight DNA methylation genes, and the formula is as follow:

**Figure 3:**
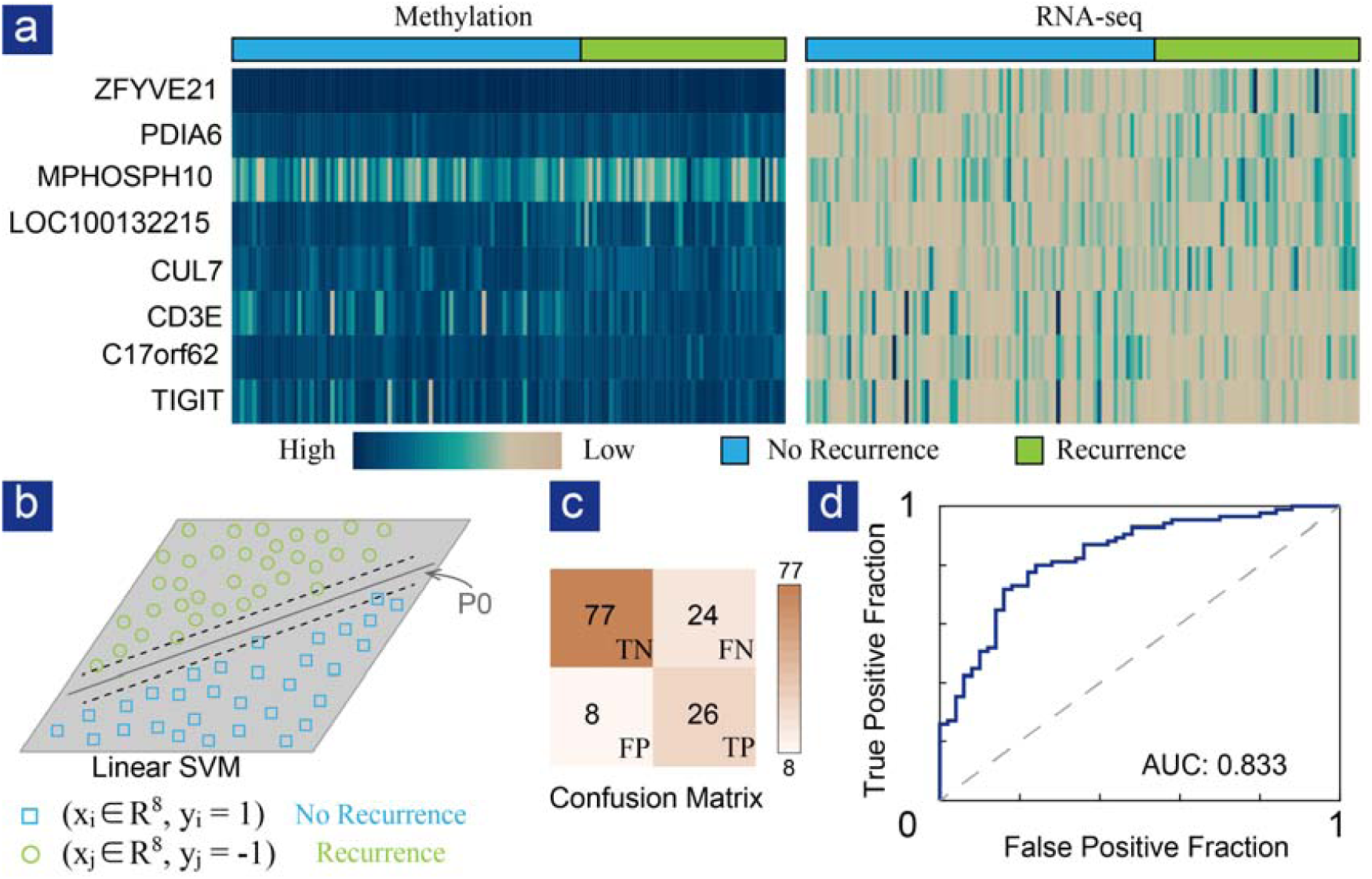
Establishment of the prediction model. (a) Heat map of 8 methylation-related genes in cervical cancer patients. (b) Schematic of the SVM algorithm. (c) The confusion matrix of the prediction model. Positive: recurrence. (d) ROC curve of the prediction model, AUC=0.833.

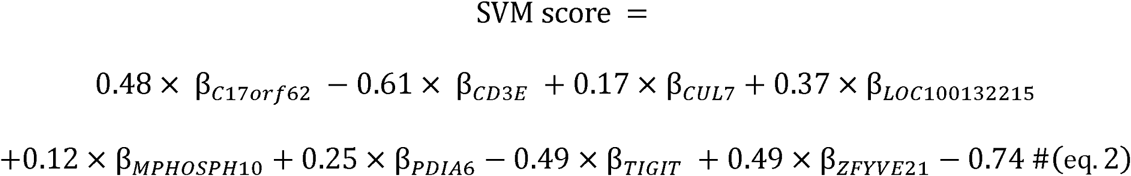

An SVM score larger than the threshold 0 is regarded as a sample with low risk, and an SVM score smaller than 0 is regarded as a sample with high risk.

### Univariate analysis and multivariate analysis

Clinical features and SVM scores of these patients were analyzed by univariate analysis using a T-test and Fisher’s exact test in R (Table 1). SVM score, FIGO stage, N stage, M stage, and pathologic grade were risk factors for recurrence (p-value < 0.05) in univariate analysis.

**Table 1:**
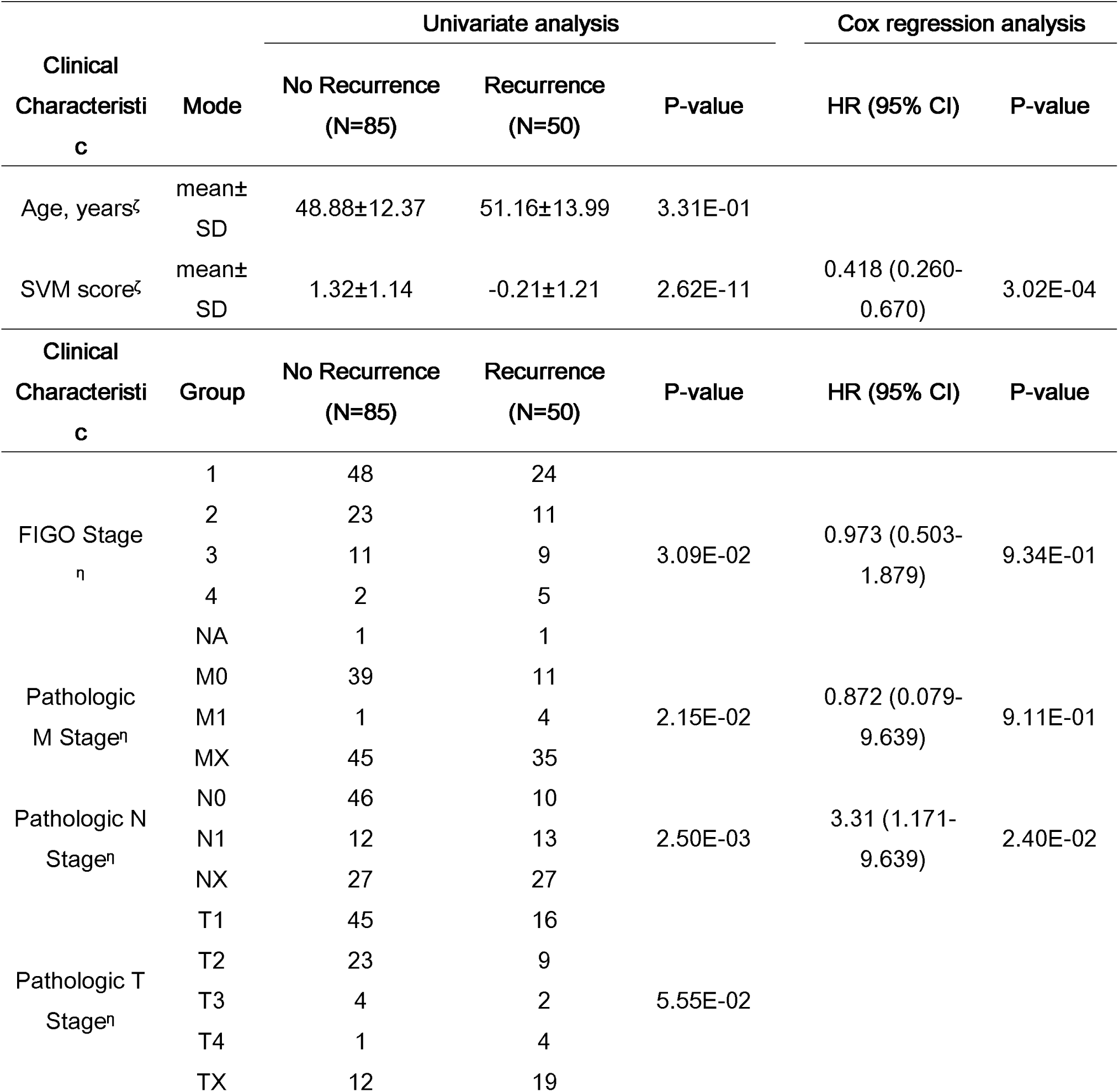
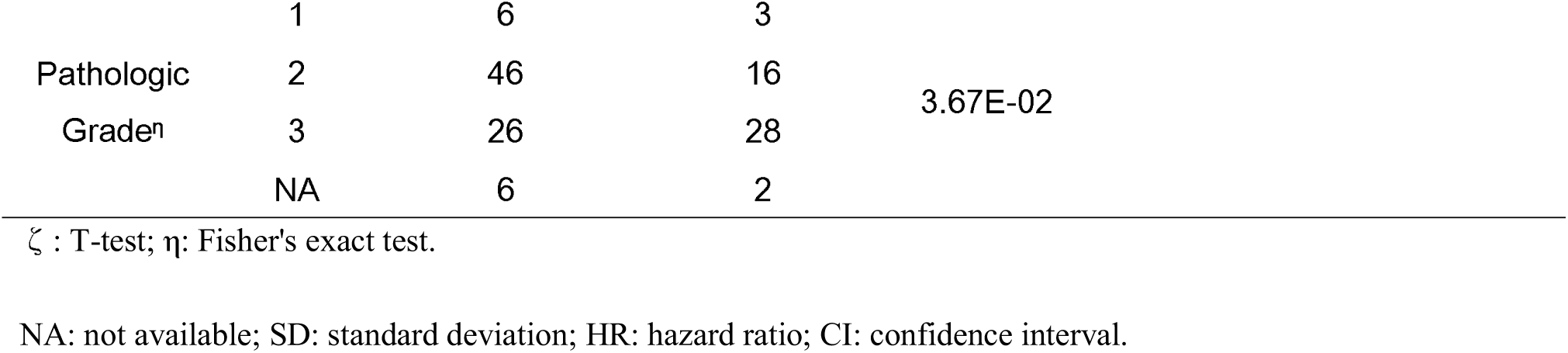
Stratification analysis for risk factors for recurrence in patients from the TCGA.

Multivariate Cox regression analysis was further conducted on these risk factors. SVM score (hazard ratio (HR) = 0.42; 95% confidence interval (CI): 0.26-0.67, p-value <0.001) and N stage (HR = 3.31; 95% CI: 1.17-9.33, p-value <0.05) were independent risk factors for recurrence(Table1).

### Visualizing the model using t-SNE

The results of our training model with t-SNE are shown in **Figure 4**. The eight-dimensional data were mapped onto a two-dimensional space by the dimensionality reduction method. Samples in each group are represented as a data point in red or blue. The results revealed that samples closer to the bottom left were more likely to be from patients in the recurrence group.

**Figure 4:**
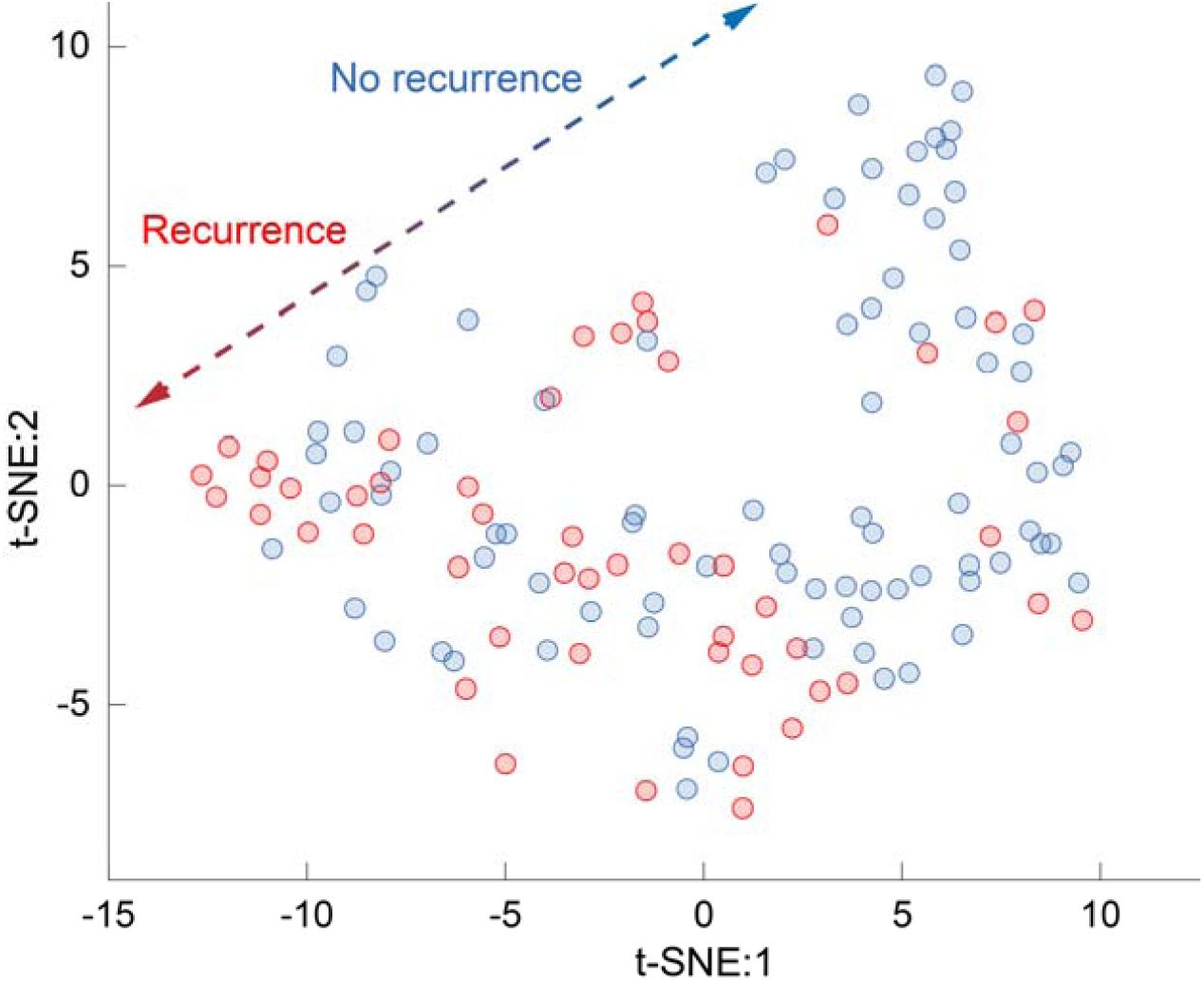
Visualization of the eight-dimensional variables with the t-SNE algorithm. Points in red represent patients with recurrence, and those in blue represent patients without recurrence.

### External validation of the GEO database

An independent external GEO cohort (GSE30759) was applied to validate the SVM-based recurrence prediction model of DNA methylation. A flowchart representation of the overall procedure is shown in Figure 5a. Three genes without the probes in the GEO dataset were substituted by the corresponding mean values in the TCGA dataset (*LOC100132215*: 0.91; *TIGIT*: 0.91; and *ZFYVE21*: 0.98) to adjust the threshold of the reduced SVM score. As shown in Figure 5b, the methylation levels of the four genes (b1: *PDIA6*; b2: *CUL7*, b3: *CD3E*, and b4: *C17orf62*) with similar distributions in two datasets formed the reduced SVM score. The mean value of *MPHOSPH10* (b5: TCGA: 0.71±0.11; GEO: 0.02±0.01) in the TCGA dataset was subtracted to further update the reduced SVM score threshold.

**Figure 5:**
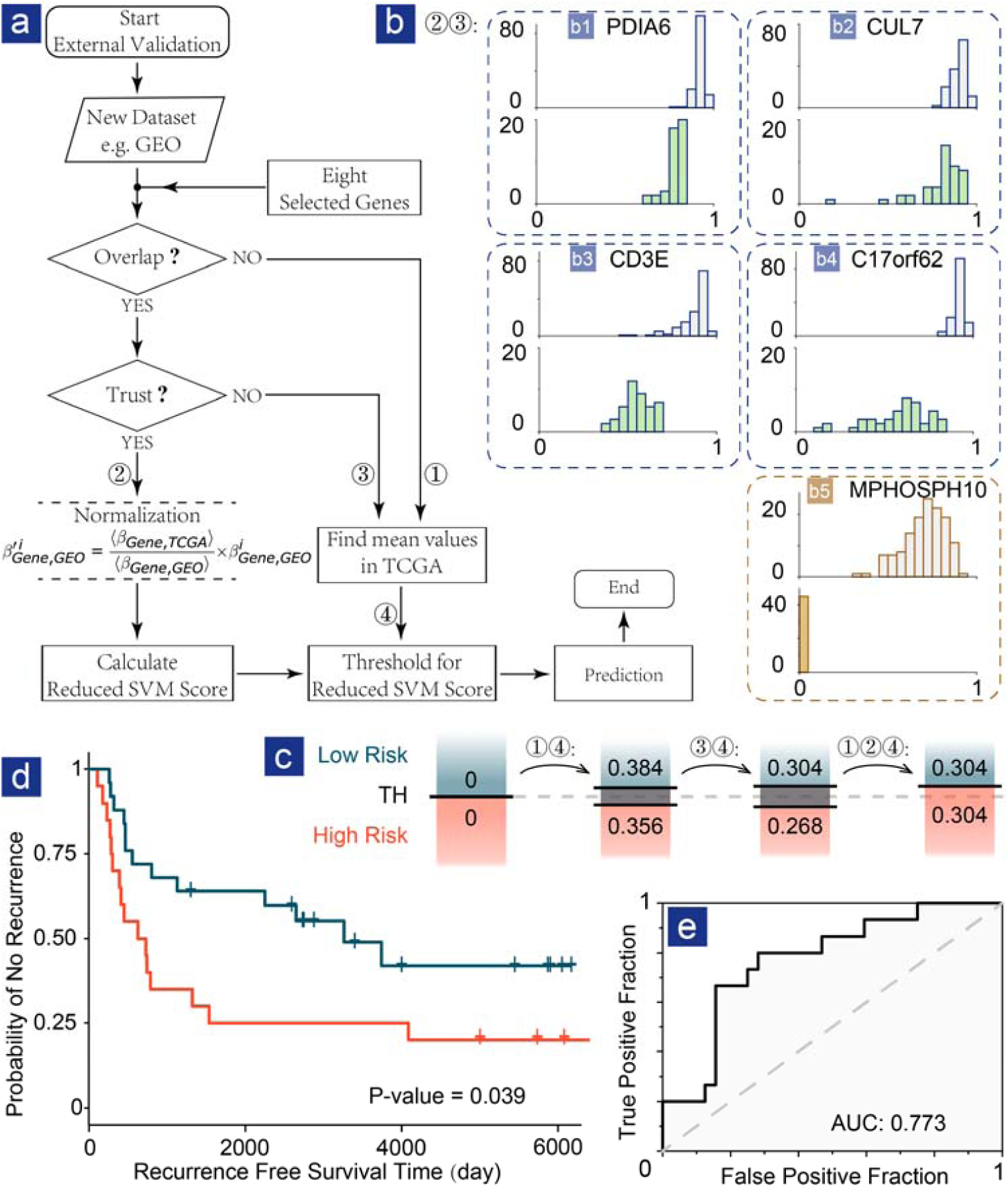
External validation of the GEO dataset. (a) Flowchart for the external validation. (b) The distribution of the methylation levels for the five genes in both the TCGA (upper) and GEO (lower) datasets. (c) Threshold shift in the procedure. (d) Survival analysis of the GEO dataset using the prediction model. (e) ROC curve of external validation with the AUC.

As shown in Figure 5c, the initial threshold of the SVM score was 0 on the basis of 8 genes. The cost of missing genes is the range of uncertainty around the threshold, where a sample is at either high or low risk. To improve the identification of high-risk patients, we considered samples within the same region as those at high risk. Therefore, the reduced SVM score formula for the GEO dataset is as follows:

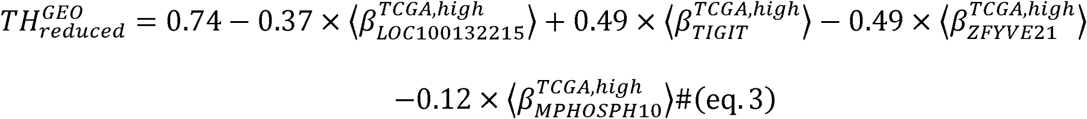

In the above expression, “high” stands for all samples at high risk, and ⟨… ⟩ indicates the average. The reduced SVM score threshold for the four genes in the GEO dataset, 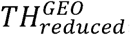, equals 0.304, instead of the initial threshold 0 for the eight genes in the TCGA dataset. The β values of the four genes (*C17orf62, CD3E, CUL7*, and *PDIA6*) from the GEO dataset then shifted the mean values to one in the TCGA dataset, avoiding bias.

The recurrence-free survival was significantly longer in the low-risk group 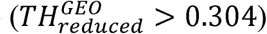 than in the high-risk group 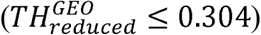 in the recurrence curve (p-value=0.039; Figure 5d). ROC curve analysis was also performed on the prediction result (Figure 5e), and the AUC was 0.773.

### GO and KEGG analyses

To research the biological function of DMGs, DAVID was used to perform GO analysis for the MFs, BPs and CCs. The MFs (p-value<0.05; **Figure 6**a) were enriched in protein binding, poly(A) RNA binding, protein homodimerization activity, and GTPase activator activity. The BPs (p-value <0.05; **Figure 6b**) were enriched in the regulation of transcription, signal transduction, and positive regulation of GTPase activity immune response. The CCs (p-value <0.05; **Figure 6c**) were enriched in the plasma membrane, cytosol, and nucleoplasm membrane. The results of KEGG analysis (**Figure 6d**) suggested that the DMGs were enriched in human T-lymphotropic virus 1 (HTLV-1) infection, cytokine-cytokine receptor interaction, T-cell receptor signaling pathway, and natural killer cell mediated cytotoxicity.

**Figure 6:**
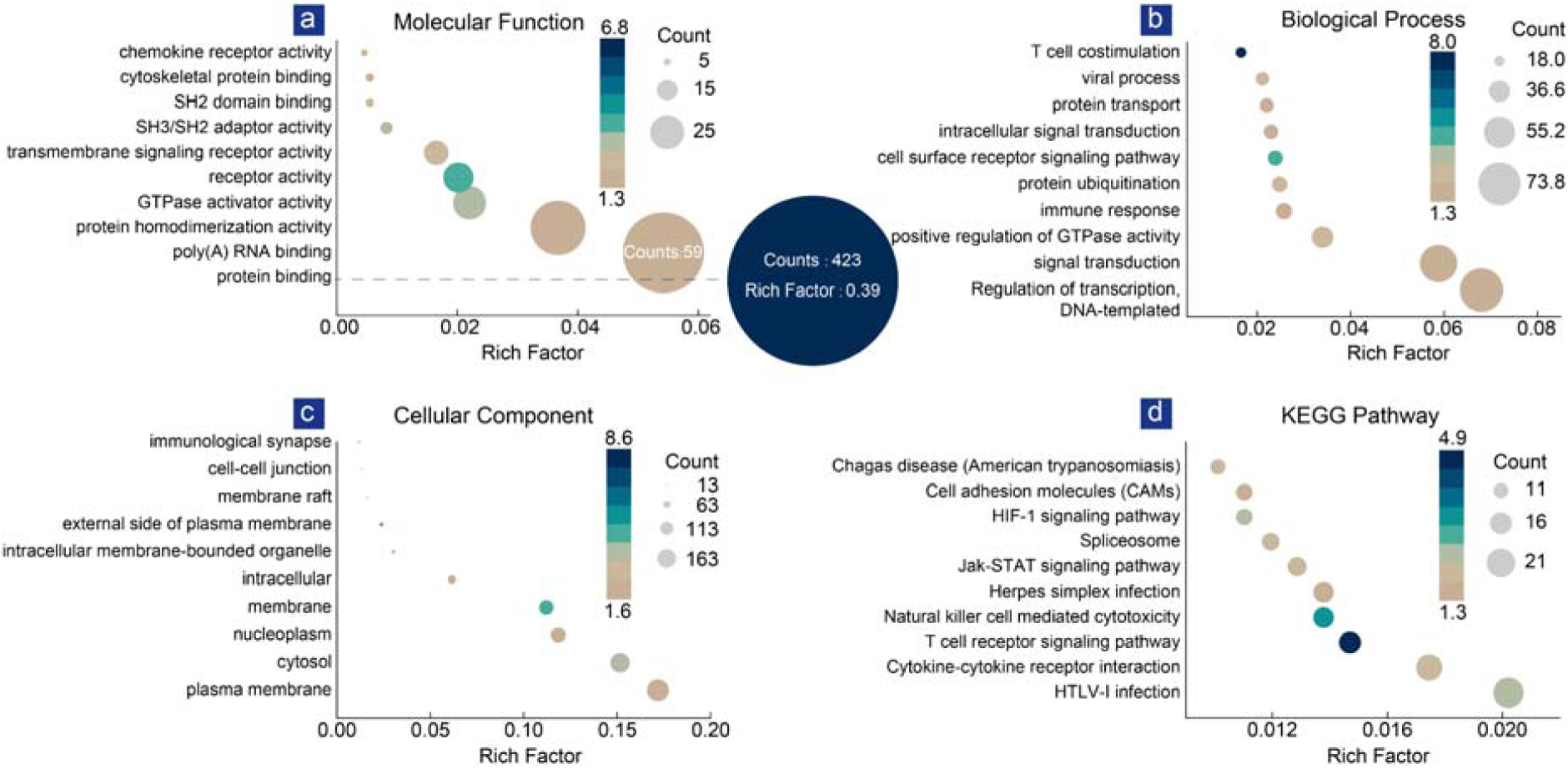
The results of GO and KEGG pathway analyses of DMGs in cervical cancer. (a) The GO terms in the MF; (b) BP; (c) CC; (d) In the results of KEGG pathway analysis, the x-axis represents the rich factor; different colors represent the −log10(p-value) and the size of the point represents the number of genes.

## Discussion

DNA methylation is a major epigenetic mechanism that plays important roles in various biological processes, such as the regulation of gene expression [31], cell differentiation [32], and inflammation [33]. DNA methylation generally occurs in CpG sites, which are unmethylated or hypomethylated in normal cells [34]. Hypermethylation of these CpG sites may silence tumor suppressor genes and lead to carcinogenesis [35]; therefore, the identification of these abnormally methylated genes contributes to the diagnosis, prediction of prognosis and selection of cancer treatment.

In this study, we performed a comprehensive screen for cervical cancer patients from the TCGA dataset by comparing patients with recurrence and without recurrence. We used the SVM machine learning algorithm along with the generated SVM scores to establish the prediction model. The SVM score has been proven to be an independent risk factor for recurrence by Cox regression analysis. In contrast, FIGO, T and M stage were not found to be independent risk factors for recurrence, which implied that the SVM score was more closely related to tumor recurrence than the traditional tumor staging system. Furthermore, the SVM score is a robust and reliable predictor for recurrence regardless of pathological type and is independent of N stage.

The ROC curve showed that the model had a high degree of fit. Independent datasets in GEO further supported the model with survival analysis. In clinical applications, this model allows clinicians to screen patients at high risk of recurrence using eight DNA methylation biomarkers.

The eight DEGs, namely, *ZFYVE21, C17orf62, PDIA6, CUL7, CD3E, TIGIT, MPHOSPH10* and *LOC100132215*, were found to be differentially affected by DNA methylation between patients with and without recurrence. Some of these genes have been reported to be dysregulated in cancer or other diseases; *ZFYVE21* is located on chromosome 14 and is associated with the metastasis of colorectal carcinoma [36] and malignancy of renal cell carcinoma [37]; *C17orf62* regulates the NADPH oxidase of phagocytes and contributes to the cause of chronic granulomatous disease [38]; *CD3E* is part of the group of immunoreceptors with tyrosine-based activation motif (ITAMs) in the T-cell receptor (TCR) signal-triggering module affecting overall survival (OS) of patients with several cancer types, including cervical cancer [39]; *TIGIT* is expressed on the surface of T and natural killer cells and is associated with the reduction in NK-cell–mediated cytotoxicity and the increase in regulatory T-cell suppression [39]; *CUL7* is an oncogenic gene that promotes cancer cell survival by promoting caspase-8 ubiquitination [39]; *LOC100132215* is correlated with gene expression in cancerous breast tissues [40]; *MPHOSPH10* is involved in ribosomal RNA (rRNA) processing during mitosis [41]; *PDIA* [40] promotes the proliferation and growth of various types of human cancer cells through activating the Wnt/β-catenin signaling pathway [39]. The role of most of these genes in cervical cancer has not been revealed, and further research is needed to determine their biological functions and mechanisms.

## Limitations

There were three main limitations in this study. First, to distinguish cervical cancer patients with and without recurrence, we excluded patients without recurrence who had a short follow-up time, which led to a decrease in the number of samples. As the follow-up time was not long enough, we included the patients who had no recurrence within two years into the no recurrence group, which may cause the deviation to some extent. Second, all the genes used for modeling methylation from the TCGA were not in the external validation datasets from GEO due to the different chips used; therefore, we had to assume the GEO dataset has the same distribution as the TCGA dataset for the three genes *LOC100132215, TIGIT*, and *ZFYVE21*. In addition, the histogram (Figure 5b) shows a large difference in the *MPHOSPH10* gene between the TCGA and GEO datasets. We calculated the average β value in the TCGA dataset and used it in the validation set. Third, further experimental studies on these genes are still needed to clarify their functions and mechanisms in cervical cancer.

## Conclusions

In this study, we identified eight recurrence-related DNA methylation biomarkers in cervical cancer through a comprehensive analysis and established a recurrence prediction model using machine learning. This work offers a preliminary but effective approach to predict cancer recurrence with a few biomarkers,which may help clinicians identify high-risk patients and implement active treatment.

## Data Availability

Not applicable.

## Data Availability

All the analyses and preparation codes, along with the related data files, will be collected and attached in the repository https://github.com/zfeng2019/8dna. The TCGA Cervical Squamous Cell Carcinoma and Endocervical Adenocarcinoma (TCGA-CESC) data were downloaded on January 28, 2016, from the Broad Institute FireBrowse Data Portal (www.firebrowse.org). The GEO dataset is available at http://www.ncbi.nlm.nih.gov/geo with the accession number GSE30759.

## Authors’ contributions

J.-H. M. and Z. F. designed the study; J.-H. M., Y. H. and L.-Y. L. carried out the analysis; J.-H. M. and Z. F. wrote the manuscript. L.-Y. L. and Z. F. visualized the analysis. All authors have read and approved the final manuscript.

## Supplementary Material

### Statistical analysis of the histopathological types

Squamous cell carcinoma and adenocarcinoma are two major histologic types of cervical cancer [1, 2]. In 135 cervical cancer samples in the TCGA database, 116 cases were squamous cell carcinoma,18 cases were adenocarcinoma and 1 case were adenosquamous cell carcinoma respectively.

Univariate analysis was performed among histological types. The result showed that there was no statistically significant difference for recurrence rate (0.379 vs 0.313 vs 1, p-value = 0.2882, Fisher’s Test) and for SVM score (0.739±1.405 vs 0.783±1.335 vs 0.022, p-value = 0.8999, T-test). Therefore, we combined patient samples with different histologic types, since histologic types of cervical cancer were not associated with the recurrence and SVM score.

We further inspected on the predominated squamous type. Univariate analysis and Cox regression analysis were conducted in **Table.S1**. The result showed that SVM score was still the independent risk factor for recurrence and T stage, instead of the N stage, became the independent risk factor for recurrence.

### Statistical analysis of the N stage

The distribution of the N stage in 135 samples is N0: 56; N1: 25; Nx: 54. The unknown N stage (Nx) accounts for nearly half of the total number of samples. To exclude the impact of Nx, we omitted samples with the Nx and performed univariate analysis and Cox regression analysis on the remaining 81 samples in **Table.S2**. The result showed that SVM score and N stage were still independent risk factors for recurrence.

We also deleted N stage and discovered that SVM score became the only independent risk factor for recurrence (**Table.S3**).

---

**Table. S 1.** Univariate and Cox regression analysis for squamous cell carcinomas only.

| Clinical Char. | Mode | Univariate analysis |  |  | Cox regression analysis |  |
| --- | --- | --- | --- | --- | --- | --- |
|  |  | No Recur.<br>(N=72) | Recur.<br>(N=44) | P-value | HR (95% CI) | P-value |
| Age, yearsζ | mean±<br>SD | 48.88±12.77 | 52.27±<br>13.99 | 2.00E-01 | | |
| SVM scoreζ | mean±<br>SD | 1.32±1.19 | -0.21±1.21 | 2.35E-09 | 0.437 (0.282-0.675) | 1.91E-04 |
| Clinical Char. | Group | No Recur.<br>(N=72) | Recur.<br>(N=44) | P-value |  |  |
| FIGO Stageη | 1 | 37 | 20 | 1.89E-01 | | |
|  | 2 | 22 | 10 |  |  |  |
|  | 3 | 11 | 9 |  |  |  |
|  | 4 | 1 | 4 |  |  |  |
|  | NA | 1 | 0 |  |  |  |
| Pathologic Mη | M0 | 36 | 10 | 1.17E-02 | 1.075 (0.522-2.216) | 8.44E-01 |
|  | M1 | 0 | 3 |  |  |  |
|  | MX | 25 | 15 |  |  |  |
| Pathologic Nη | N0 | 35 | 9 | 2.63E-02 | 0.953 (0.433-2.098) | 9.04E-01 |
|  | N1 | 11 | 12 |  |  |  |
|  | NX | 16 | 10 |  |  |  |
| Pathologic Tη | T1 | 34 | 13 | 3.17E-02 | 1.537(1.018-2.317) | 4.11E-02 |
|  | T2 | 21 | 8 |  |  |  |
|  | T3 | 4 | 1 |  |  |  |
|  | T4 | 1 | 4 |  |  |  |
|  | TX | 2 | 5 |  |  |  |
| Pathologic<br>Gradeη | 1 | 4 | 2 | 1.88E-01 | | |
|  | 2 | 38 | 15 |  |  |  |
|  | 3 | 24 | 24 |  |  |  |
|  | NA | 5 | 2 |  |  |  |

$\zeta$ : T test; $\eta$ : Fisher's exact test.
NA: not available; SD: standard deviation; HR: hazard ratio; CI: confidence interval.

**Table. S 2.**
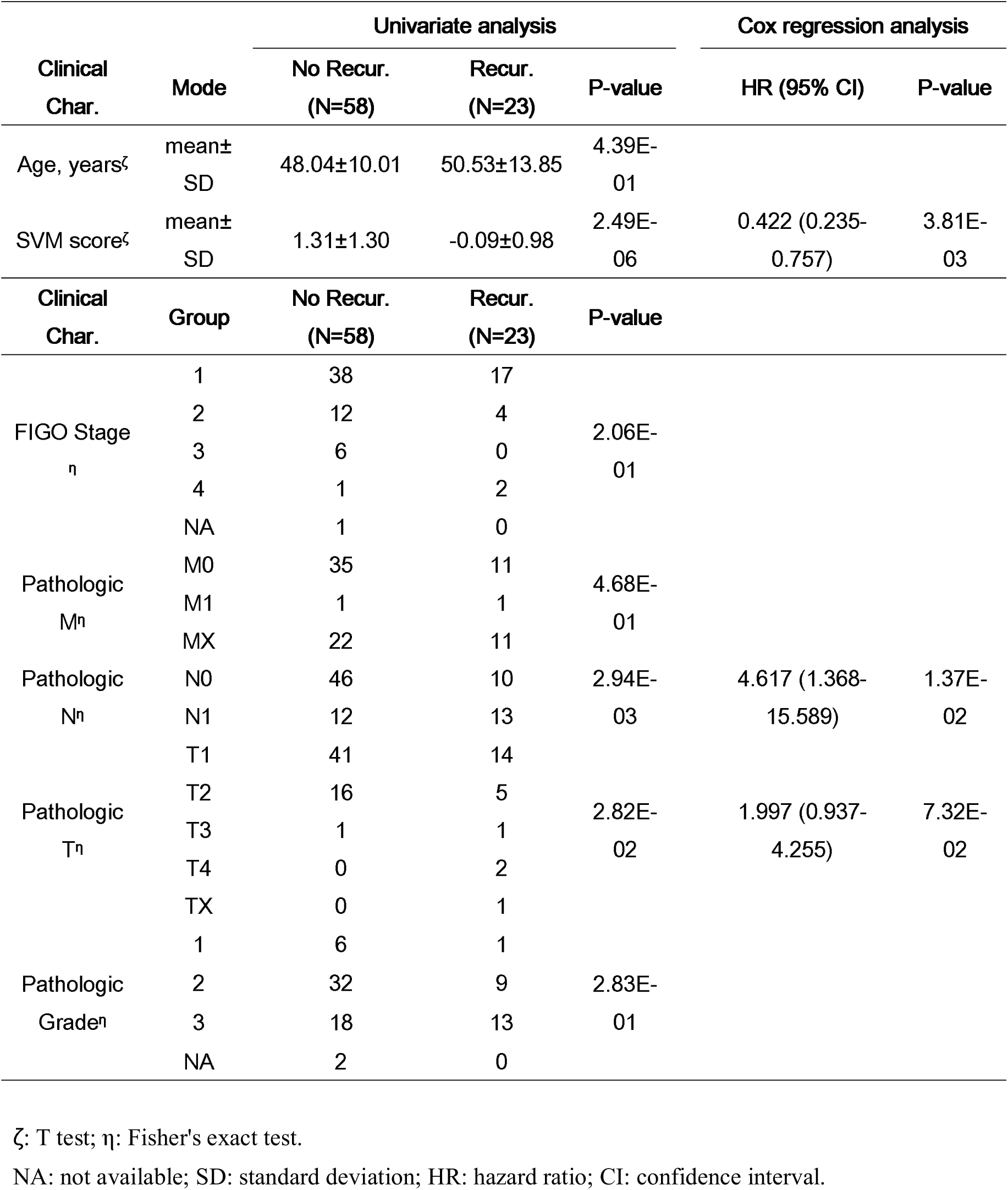
Univariate and Cox regression analysis irrespective of samples with the Nx.

**Table. S 3.** Univariate and Cox regression analysis without the N stage.

| Clinical | Mode | Univariate analysis |  |  | Cox regression analysis |  |
| --- | --- | --- | --- | --- | --- | --- |
|  |  | No Recur. | Recur. | P-value | HR (95% CI) | P-value |

| Char. |  | (N=85) | (N=50) |  |  |  |
| --- | --- | --- | --- | --- | --- | --- |
| Age, years<sup>ζ</sup> | mean±SD | 48.88±12.37 | 51.16±13.99 | 3.31E-01 |  |  |
| SVM score<sup>ζ</sup> | mean±SD | 1.32±1.14 | -0.21±1.21 | 2.62E-11 | 0.452 (0.302-0.676) | 1.11E-04 |
| Clinical Char. | Group | No Recur.<br>(N=85) | Recur.<br>(N=50) | P-value |  |  |
| FIGO Stage<sup>η</sup> | 1 | 48 | 24 |  |  |  |
|  | 2 | 23 | 11 |  |  |  |
|  | 3 | 11 | 9 | 3.09E-02 | 0.777 (0.422-1.433) | 4.19E-01 |
|  | 4 | 2 | 5 |  |  |  |
|  | NA | 1 | 1 |  |  |  |
| Pathologic M<sup>η</sup> | M0 | 39 | 11 |  |  |  |
|  | M1 | 1 | 4 | 2.15E-02 | 0.767 (0.454-1.295) | 3.20E-01 |
|  | MX | 45 | 35 |  |  |  |
| Pathologic T<sup>η</sup> | T1 | 45 | 16 |  |  |  |
|  | T2 | 23 | 9 |  |  |  |
|  | T3 | 4 | 2 | 5.55E-02 |  |  |
|  | T4 | 1 | 4 |  |  |  |
|  | TX | 12 | 19 |  |  |  |
| Pathologic Grade<sup>η</sup> | 1 | 6 | 3 |  |  |  |
|  | 2 | 46 | 16 | 3.67E-02 |  |  |
|  | 3 | 26 | 28 |  |  |  |
|  | NA | 6 | 2 |  |  |  |

ζ: T test; η: Fisher's exact test.
NA: not available; SD: standard deviation; HR: hazard ratio; CI: confidence interval.

